# Identification of established and novel extracellular matrix components in glioblastoma as targets for angiogenesis and prognosis

**DOI:** 10.1101/2024.02.28.24303381

**Authors:** Lucas Cunha Barbosa, Gabriel Cardoso Machado, Manoela Heringer, Valéria Pereira Ferrer

**Affiliations:** Graduation Program of Pathological Anatomy, Faculty of Medicine, Federal University of Rio de Janeiro; Brain’s Biomedicine Lab, Paulo Niemeyer State Brain Institute, Rio de Janeiro, Brazil; Laboratory of Cellular and Molecular Biology of Tumors, Department of Cellular and Molecular Biology, Institute of Biology, Fluminense Federal University, Niteroi, Brazil

**Keywords:** extracellular matrix, glioblastoma, endothelial cells, biomarkers, prognosis

## Abstract

Glioblastomas (GBM) are aggressive tumors known for their heterogeneity, rapid proliferation, treatment resistance, and extensive vasculature. Angiogenesis, the formation of new vessels, involves endothelial cell (EC) migration and proliferation. Various extracellular matrix (ECM) molecules regulate EC survival, migration, and proliferation. Culturing human brain EC (HBMEC) on GBM-derived ECM revealed a decrease in EC numbers compared to controls. Through *in silico* analysis, we explored ECM gene expression differences between GBM and brain normal glia cells and the impact of GBM microenvironment on EC ECM transcripts. ECM molecules such as collagen alpha chains (*COL4A1*, *COL4A2*, p < 0.0001); laminin alpha (*LAMA4*), beta (*LAMB2*), and gamma (*LAMC1*) chains (p < 0.0005); neurocan (*NCAN*), brevican (*BCAN*) and versican (*VCAN*) (p < 0.0005); hyaluronan synthase (*HAS*) 2 and metalloprotease (*MMP*) 2 (p < 0.005); MMP inhibitors (*TIMP1*-*4*, p < 0.0005), transforming growth factor beta-1 (*TGFB1*) and integrin alpha (*ITGA3*/5) (p < 0.05) and beta (*ITGB1*, p < 0.0005) chains showed increased expression in GBM. Additionally, GBM-influenced EC exhibited elevated expression of *COL5A3*, *COL6A1*, *COL22A1* and *COL27A1* (p < 0.01); *LAMA1*, *LAMB1* (p < 0.001); fibulins (*FBLN1*/*2*, p < 0.01); *MMP9*, *HAS1*, *ITGA3*, *TGFB1*, and wingless-related integration site 9B (*WNT9B*) (p < 0.01) compared to normal EC. Some of these molecules: *COL5A1*/*3*, *COL6A1*, *COL22/27A1*, *FBLN1/2*, *ITGA3/5*, *ITGB1* and *LAMA1/B1* (p < 0.01); *NCAN*, *HAS1*, *MMP2/9*, *TIMP1/2* and *TGFB1* (p < 0.05) correlated with GBM patient survival. In conclusion, this study identified both established and novel ECM molecules regulating GBM angiogenesis, suggesting *NCAN* and *COL27A1* are new potential prognostic biomarkers for GBM.

## Introduction

Adult diffuse gliomas are brain tumors that originate from glial cells. These tumors are currently categorized into three subtypes: (1) astrocytoma with mutations in the isocitrate dehydrogenase (IDH) gene; (2) oligodendroglioma with IDH mutations and chromosome 1p/19q-codeletion; (3) GBM, which is the most aggressive type of glioma, IDH wild-type followed by at least one of these characteristics: amplification of the EGFR gene, mutation in the TERT promoter, and/or the concurrent gain of chromosome 7 and loss of chromosome 10 (+7/−10), necrosis and microvascular proliferation [1, 2]. The median survival time after GBM diagnosis is still approximately 15 months, and the 5-year overall survival rate is only 6.8% [3].

GBM growth depends on the formation of new blood vessels, and GBM is one of the most vascularized tumors [4]. Blood vessels that develop in the primary tumor are larger than those in normal counterparts and follow a crisscrossing path, with larger and irregular lumen diameters, high permeability, and irregular branching [5]. The tumor vasculature is hyperpermeable to plasma, leading to local edema and extravascular clotting of plasma [6, 7], altered blood flow and flux of leukocytes in the tumor site [8], and the spread of tumor cells [9, 10]. The leakiness of vessels leads to large volumes of tumor tissue without blood flow and obstructs the delivery of drugs, oxygen, and nutrients, resulting in ischemia and necrotic regions within the tumor [11, 12].

Angiogenesis is a complex process involving the proliferation, migration, and differentiation of vascular EC under stimulation by specific signals [13]. In brief, the lack of oxygen in the EC stimulates the increase and release of proangiogenic growth factors such as vascular endothelial growth factor (VEGF) [14, 15], TGFB1 [16, 17], stromal-derived factor 1 (SDF-1) [18, 19], and angiopoetins [20, 21], among others [22]. These angiogenic factors bind to their receptors on the EC membrane, leading to the dissolution of the vessel wall and degradation of the EC basement membrane (BM) and the surrounding ECM [23]. Following the degradation of the BM, specific proteases such as MMP remodel ECM components [24], and a new matrix is synthesized by stromal cells, which in turn promotes the migration and proliferation of EC resulting in the formation of an endothelial tube-like structure [25]. Then, a mature vascular BM is formed around the newly formed endothelial tube, and the pericytes and smooth muscle cells surround it, resulting in a stable new vessel [13].

The brain and GBM ECM are enriched in hyaluronic acid (HA); proteoglycans such as NCAN and VCAN; and glycoproteins such as tenascin and fibronectin [26–28]. ECM molecules themselves trigger EC signaling through the activation of specific membrane receptors and modulate access to stored growth factors, assisting several processes in angiogenesis, such as EC attachment and detachment, EC survival, proliferation and migration [13, 23]. The mechanical properties of the ECM also contribute to modulating these angiogenic processes [29]. Therefore, ECM molecules can have both pro- and antiangiogenic effects [30, 31].

Certain ECM components, such as tenascin C [31–33], collagens I, IV and VI [34, 35], HA [36], laminin [37], VCAN and fibronectin [28], are known to modulate GBM angiogenesis. Given the significance of the ECM and vascularity in GBM aggressiveness, as well as the dual angiogenic effects of ECM components, we aimed to investigate the influence of the GBM ECM on the total number of brain EC and on ECM gene expression in these cells. Additionally, we evaluated the potential direct contribution of the expression of certain ECM genes to the OS of GBM patients using *in silico* analysis.

## Materials and Methods

### Cell Culture

GBM cells (GBM02) were established by Professor Vivaldo Moura-Neto’s group (Paulo Niemeyer State Brain Institute) via surgical resection of a human tumor in 2002 [38] and cordially provided for this study. The HBMEC lineage originating from the Prof. Dennis Grab Laboratory (Department of Pathology, Johns Hopkins School of Medicine) [39] was generously provided by Professor Catarina Freitas’s research group. The cells were thawed from liquid nitrogen and plated in 75 cm2 cell culture flasks containing Dulbecco’s modified Eagle’s medium/Ham’s F12 media (DMEM/F-12) supplemented with 10% fetal bovine serum (FBS). The cells were maintained in a humidified incubator with 5% CO_2_ at 37°C. The ethical committee approval by the National Commission (CONEP) is under the number 2340.

### Formation of ECM-free of cells

A total of 5x10^4^ HBMEC (control) or GBM02 cells were cultured on glass coverslips coated with 0.1% bovine gelatin in 24-well plates [40]. The cells were maintained in a humidified incubator with 5% CO_2_ at 37°C in DMEM/F12 supplemented with 10% FBS. After 72 h of cultivation, the cells were lysed using buffer containing 0.1% Triton X-100 and 0.1 M NH_4_OH in PBS at 4°C. Gentle washes with PBS were performed to eliminate cellular debris [27]. The remaining protein network, which was invisible under an optical microscope, was termed the preformed ECM-free of cells.

### HBMEC immunofluorescence and counting

HBMEC (5x10^4^) were cultivated with HBMEC ECM (control) or GBM ECM in a 24-well plate covered with 0.1% gelatin for 24 h, 48 h or 72 h. At these time points, the cells were fixed with 4% paraformaldehyde (PFA)/PBS for 20 min, permeabilized with 0.1% Triton X-100/PBS, and blocked with 5% BSA/PBS for 1 h [24]. The coverslips were then treated with 0.165 μM Alexa Fluor 488 phalloidin solution (A12379, Invitrogen) for 30 min and washed with PBS. Then, they were stained with DAPI (4’,6-diamino-2-phenylindole) at 1 μL/mL (D9542 Sigma[Aldrich®) and mounted on glass slides using Fluoromount-G®. The cells were imaged at 20x using a DMi8 advanced fluorescence microscope (Leica Microsystems, Wetzlar, Germany) and analyzed with a Leica LAS AF Lite. The images were processed, and nuclei were counted using Fiji software for ImageJ version 1.52 (Wayne Rasband, National Institutes of Health, MD, USA). The analyses were performed using three biological replicates, and statistics were performed using analysis of variance (ANOVA) in GraphPad Prism version 9.

### Bioinformatic analysis

#### Expression of ECM components in GBM cells and influence of the GBM microenvironment on the expression of ECM transcripts in EC

We retrieved expression data from GBM cells (N=865) and brain normal glial cells (N=27) by accessing the Single Cell Portal (https://singlecell.broadinstitute.org/single_cell) and specifically selected the dataset titled “Single-cell multiomics profiling of human gliomas” [41]. Additionally, a curated database of EC transcriptomics data (EndoDB, https://endotheliomics.shinyapps.io/endodb/) was used to download transcript data from the EC [42]. We selected the E-GEOD-9861, E-MTAB-5686, and E-MTAB-765 [43] studies to obtain data on 11 samples of EC from normal brains. Additionally, we chose the E-MEXP-2891 [44] study, from which we obtained data on 6 samples of EC from GBM specimens. We compared the expression of a set of ECM molecules (proteoglycans, glycoproteins, synthesizing and degrading enzymes and stored growth factors) between normal and GBM data. First, we performed the D’Agostino-Pearson normality test and then used the Mann[Whitney test for statistical analysis. The statistical analysis and generation of graphs were performed using GraphPad Prism version 9.

#### OS curves

Survival data were utilized to ascertain the final recorded day of patient survival. Comparative analyses of survival among the specified genes were performed via the Kaplan[Meier method and the log-rank (Mantel–Cox) test via GraphPad Prism version 9. We selected ECM molecules that presented interesting results in the EC expression analysis to verify the influence of their direct expression on the OS of patients with GBM. Using the transcriptome information of the Human Protein Atlas (https://www.proteinatlas.org/) [45] and The Cancer Genome Atlas (TCGA) databases (https://www.cbioportal.org/) [46, 47] (N = 153), we divided the patients into high- and low-expression groups. The best expression cutoff referred to the value that yielded the maximal difference with regard to survival between the two groups at the lowest log-rank P value and was determined at the Human Protein Atlas site. The median survival was determined as the minimal survival time at which the survival function equaled or fell below 50%.

## Results

### The GBM-derived ECM suppressed the number of HBMEC

Initially, we investigated whether the GBM ECM influenced the quantity of HBMEC at 24, 48 and 72 h. At 24 h, we observed a significant decrease in the number of HBMEC induced by the GBM ECM (** p < 0.01) compared to that induced by the gelatin coverslip control (Fig. 1A). However, at 48 h, the HBMEC counts were similar across all the tested conditions (Fig. 1B). Notably, at 72 h, compared with the HBMEC ECM, the GBM ECM continued to decrease the total quantity of HBMEC (** p < 0.01) (Fig. 1C). These results collectively demonstrate that the GBM ECM can reduce the total number of HBMEC.

**Fig. 1.**
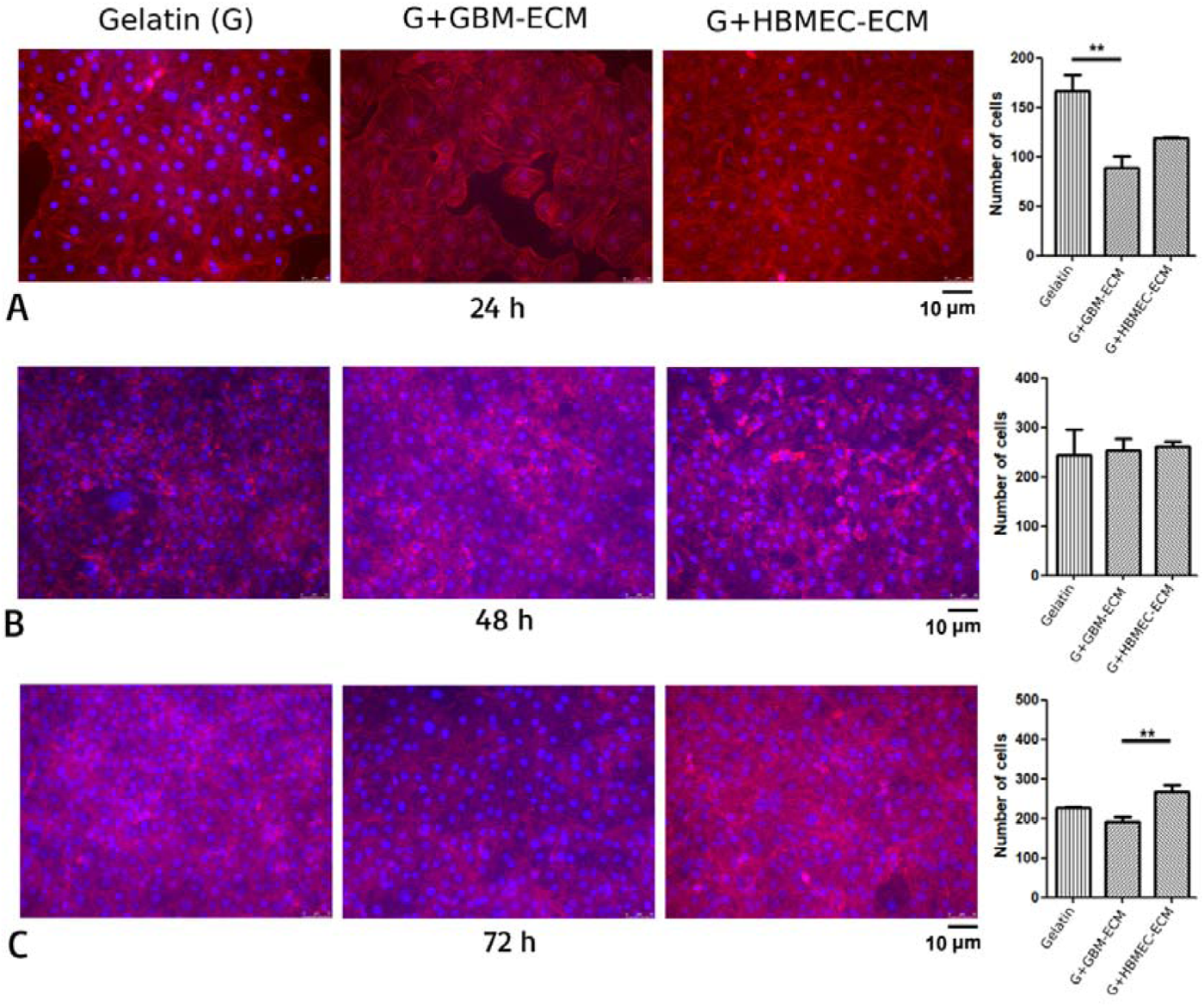
The GBM-derived ECM decreased the total number of HBMEC at 24 h and 72 h. Following 72 h of GBM02 and HBMEC cultivation on gelatin coverslips (G), a cell-free GBM-derived ECM (GBM-ECM) and HBMEC-derived ECM (HBMEC-ECM) were obtained using Triton X-100 lysis buffer. Subsequently, HBMEC were cultured under gelatin control or HBMEC-ECM or GBM-ECM conditions for (A) 24 h, (B) 48 h, or (C) 72 h. Immunofluorescence staining using phalloidin (actin, in red) and DAPI (nuclei, in blue) was performed. The nuclei were quantified using ImageJ software and statistically analyzed using ANOVA. At 24 h, the GBM ECM exhibited a reduced number of HBMEC compared to that in the gelatin coverslip control (** p < 0.01). Similarly, at 72 h, the number of HBMEC in the GBM ECM was lower than that in the HBMEC ECM control (** p < 0.01). At 48 h, there were no significant differences among the conditions analyzed.

### The expression of several ECM and ECM-related genes is increased in GBM cells compared to that in normal glial cells

Since the ECM derived from GBM cells reduced the number of HBMEC, we investigated which ECM or ECM-related molecules exhibited increased expression in GBM cells compared to that in normal glia. We utilized the single-cell portal to obtain single-cell transcript information. Our observations revealed increased expression of several ECM molecules in GBM cells (Fig. 2): *COL4A1*, *COL4A2*, *LAMC1*, *LAMB2*, *VCAN* and *BCAN* (**** p < 0.0001), as well as *LAMA4* and *NCAN* (*** p < 0.0005). Additionally, we noted increased expression of the following ECM-modifying proteins (Fig. 3): *MMP2*, *TIMP1/2/4* (**** p < 0.0001), *TIMP3* and *ITGB1* (*** p < 0.0005), *HAS2* (** p < 0.005), *ITGA3* (** p < 0.005), *ITGA5* (* p < 0.05), and *TGFB1* (* p < 0.05). Conversely, there were no significant differences (ns = nonsignificant) in the expression levels of *COL1A1*, *COL4A6*, *LAMA1*/*2*, *LAMC3*, *FBLN1*/*5*, *HAS3*, *MMP9*, *ITGA1*, or *ITGB2/3/5* (data not shown). Therefore, our data revealed that GBM cells predominantly upregulated the expression of *VCAN*, *BCAN*, *NCAN*, certain subtypes of collagens, laminins, and integrin genes, *HAS2*, MMP inhibitors and *TGFB1*.

**Fig. 2.**
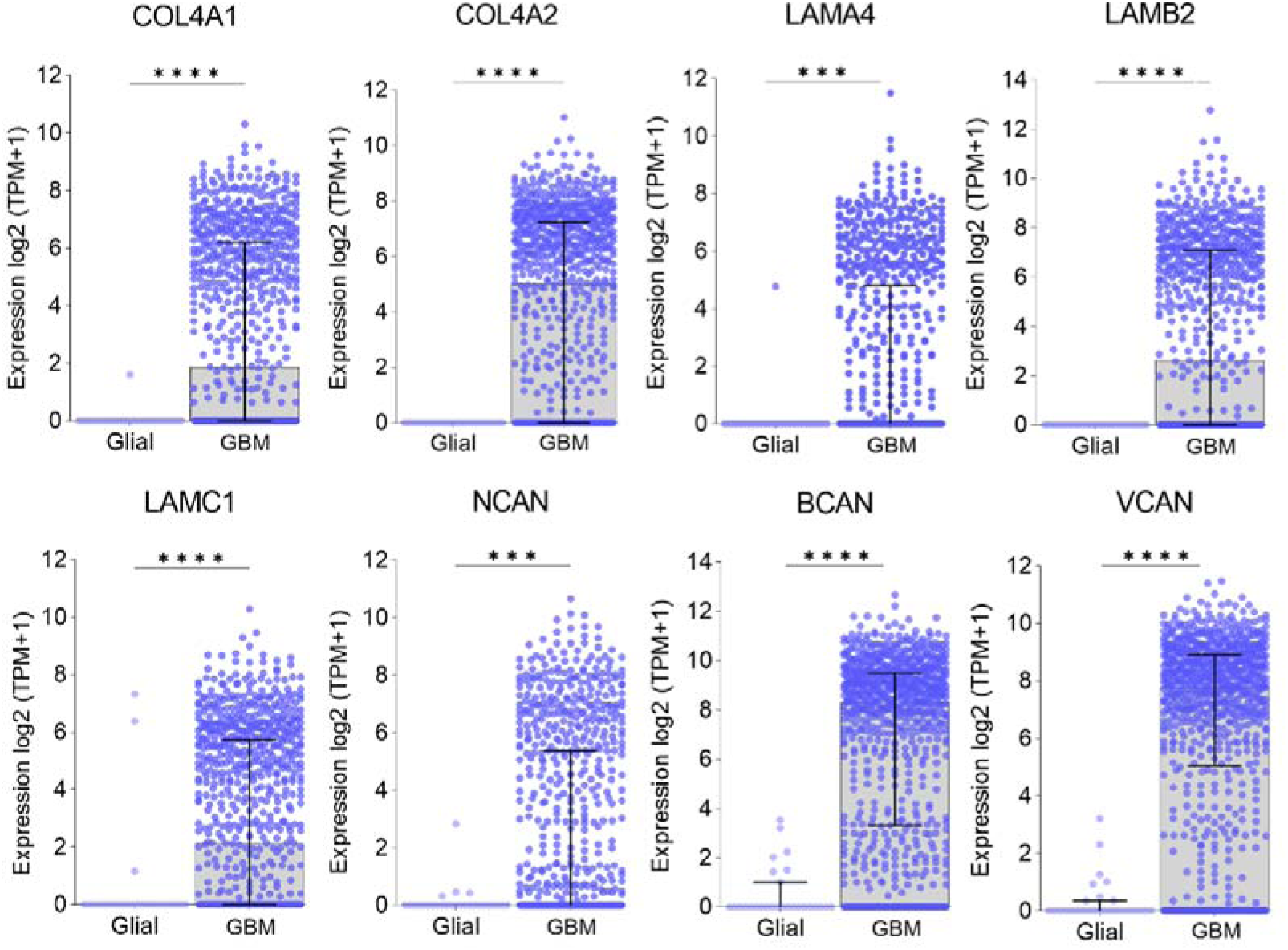
Compared with normal glia, GBM cells exhibit increased expression of certain ECM molecules. We analyzed which ECM components were increased in GBM cells compared to normal glial cells using the single-cell portal database. Data from 27 brain normal glial cells and 865 GBM cells were obtained. Among proteoglycans and glycoproteins, we observed increased expression of collagens (*COL4A1* and *COL4A2*), laminins (*LAMA4*, *LAMB2*, *LAMC1*) and lecticans (*NCAN*, *BCAN* and *VCAN*). The D’Agostino-Pearson and Mann[Whitney tests were performed for statistical analysis (*** p < 0.001, **** p < 0.0001).

**Fig. 3.**
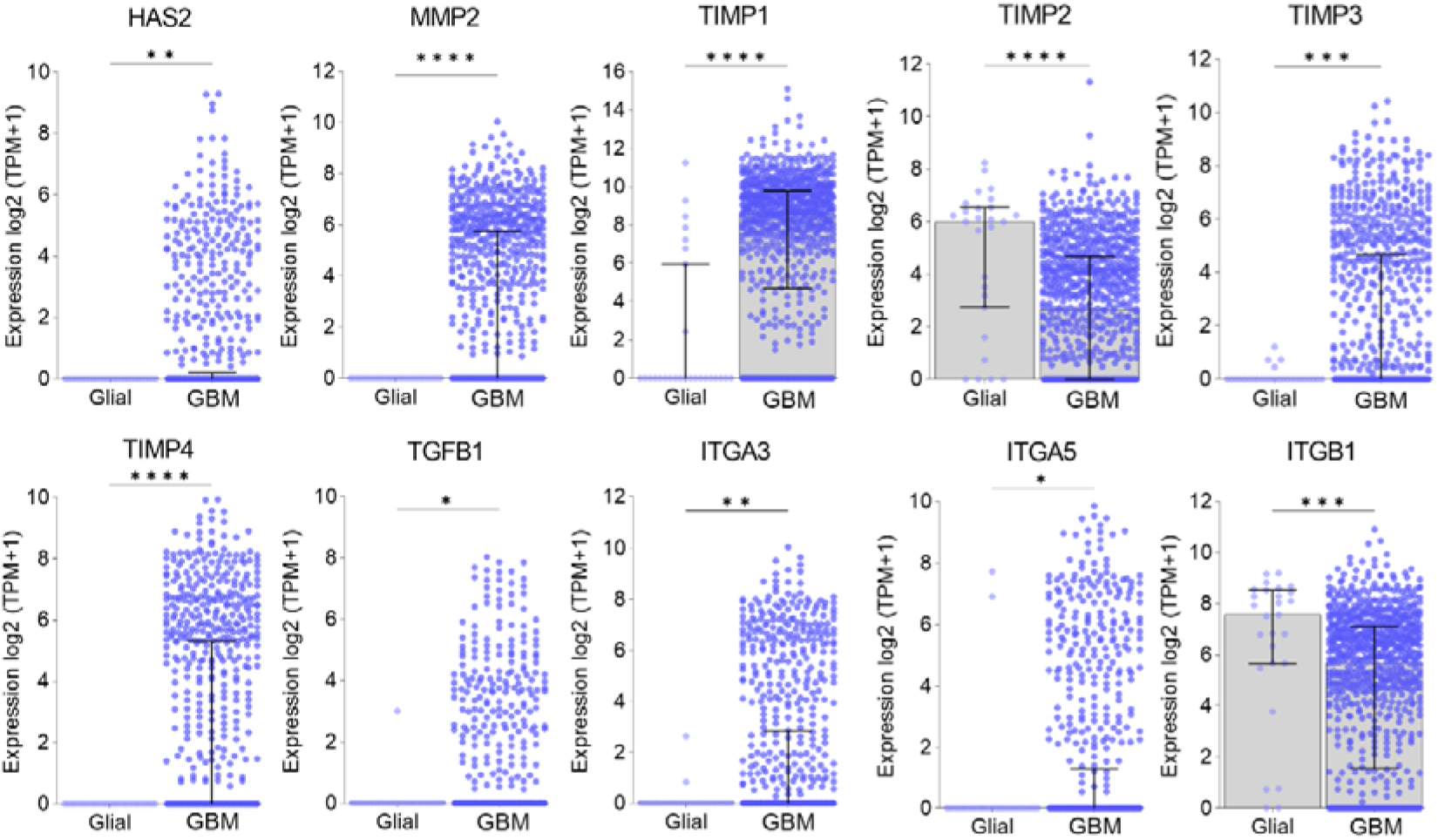
GBM cells exhibited increased expression of ECM-modifying proteins, integrin alpha chains and *TGFB1*. The expression of ECM-related proteins in normal glial brain cells (N=27) and GBM cells (N=865) was analyzed via the single-cell portal of the transcriptome database. The D’Agostino-Pearson and Mann[Whitney tests were used for statistical analysis (* p < 0.05, ** p < 0.01, *** p < 0.001, **** p < 0.0001). We observed increased expression of the ECM-modifying enzymes *HAS2* and *MMP2*, the inhibitor *TIMP1*-*4*, the cell membrane proteins *ITGA3*/*5* and *ITGB1*, and the stored growth factor *TGFB1*.

### EC exhibit increased expression of numerous ECM-related genes under the influence of the GBM microenvironment

We sought to investigate whether the GBM microenvironment alters the EC transcriptome profile of ECM-related molecules. To accomplish this, we utilized the EndoDB database. We observed that the GBM microenvironment increased the expression of several ECM genes in EC (Fig. 4): *COL5A3*, *COL22A1*, *COL27A1*, *LAMA1*, *LAMB1*, and *FBLN1* (*** p < 0.001) and *COL6A1* and *FBLN2* (** p < 0.01). However, there were no significant differences in the expression of *COL5A1* or *NCAN* in EC. We observed that EC from GBM specimens exhibited increased expression of the ECM-stored growth factors *TGFB1* and *WNT9B* (** p < 0.01), as well as the ECM-regulatory proteins *MMP9* (*** p < 0.001), *HAS1* and *ITGA3* (** p < 0.01) (Fig. 5). However, there were no significant differences in the expression of *TIMP4* between normal and GBM EC. Overall, our findings demonstrate that GBM upregulates the expression of several key ECM components in EC.

**Fig. 4.**
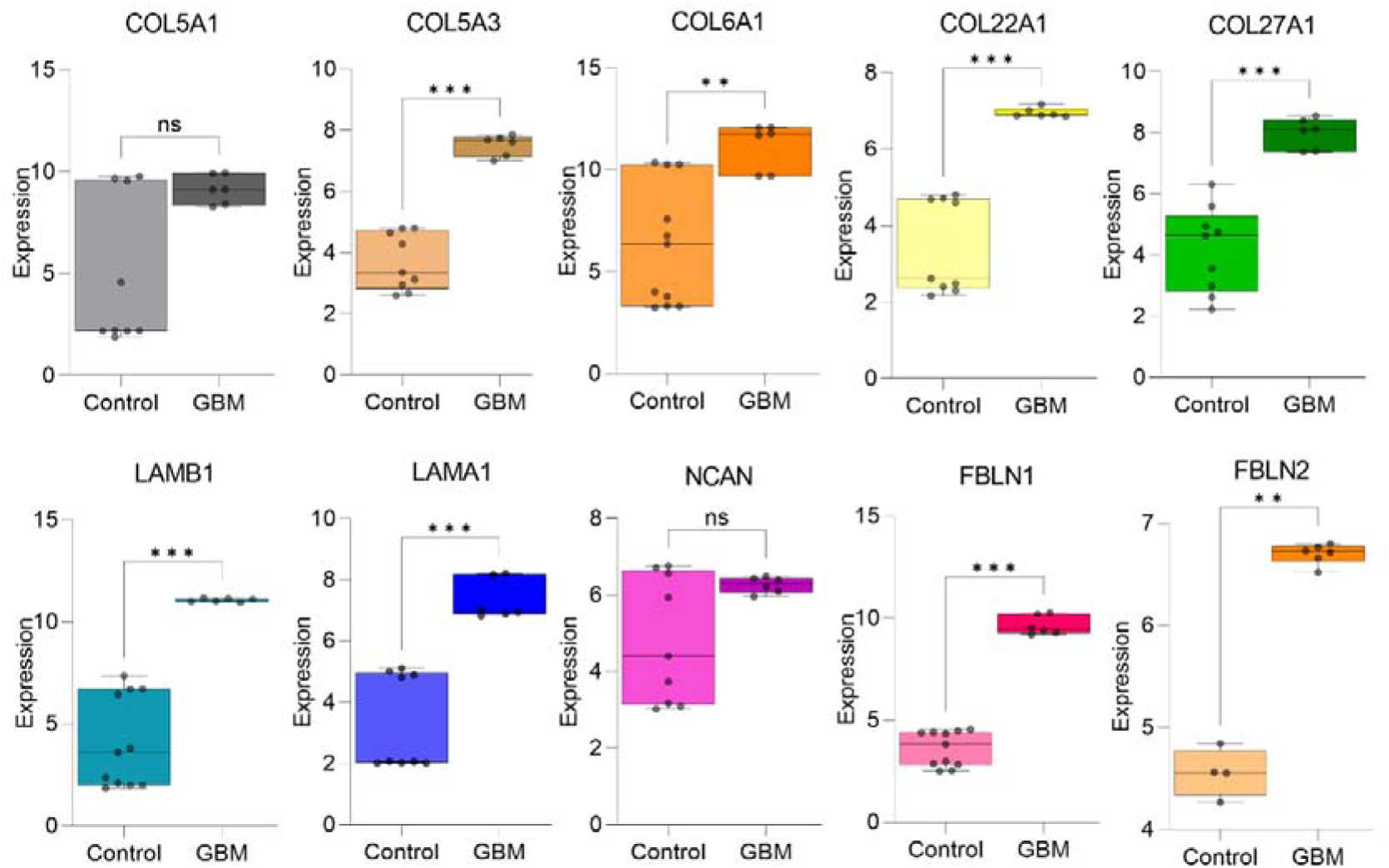
Under the influence of the GBM microenvironment, EC exhibit increased expression of multiple ECM molecules. Using data obtained from the EndoDB database, we analyzed whether the GBM microenvironment would alter the EC ECM transcriptome compared to that of normal brain EC. We obtained data from 11 samples of EC from normal brains and 6 samples from GBM EC. We analyzed the expression data of proteoglycans and glycoproteins. Statistics were performed using the D’Agostino-Pearson and Mann[Whitney tests. We observed increased expression of several ECM genes, including collagens (*COL5A3*, *COL6A1*, *COL22A1*, and *COL27A1*), laminins (*LAMA1* and *LAMB1*), and fibulins (*FBLN1* and *FBLN2*), in EC under GBM conditions. *COL5A1* and *NCAN* showed no significant (ns) differences compared to the control. (** p < 0.01, *** p < 0.001).

**Fig. 5.**
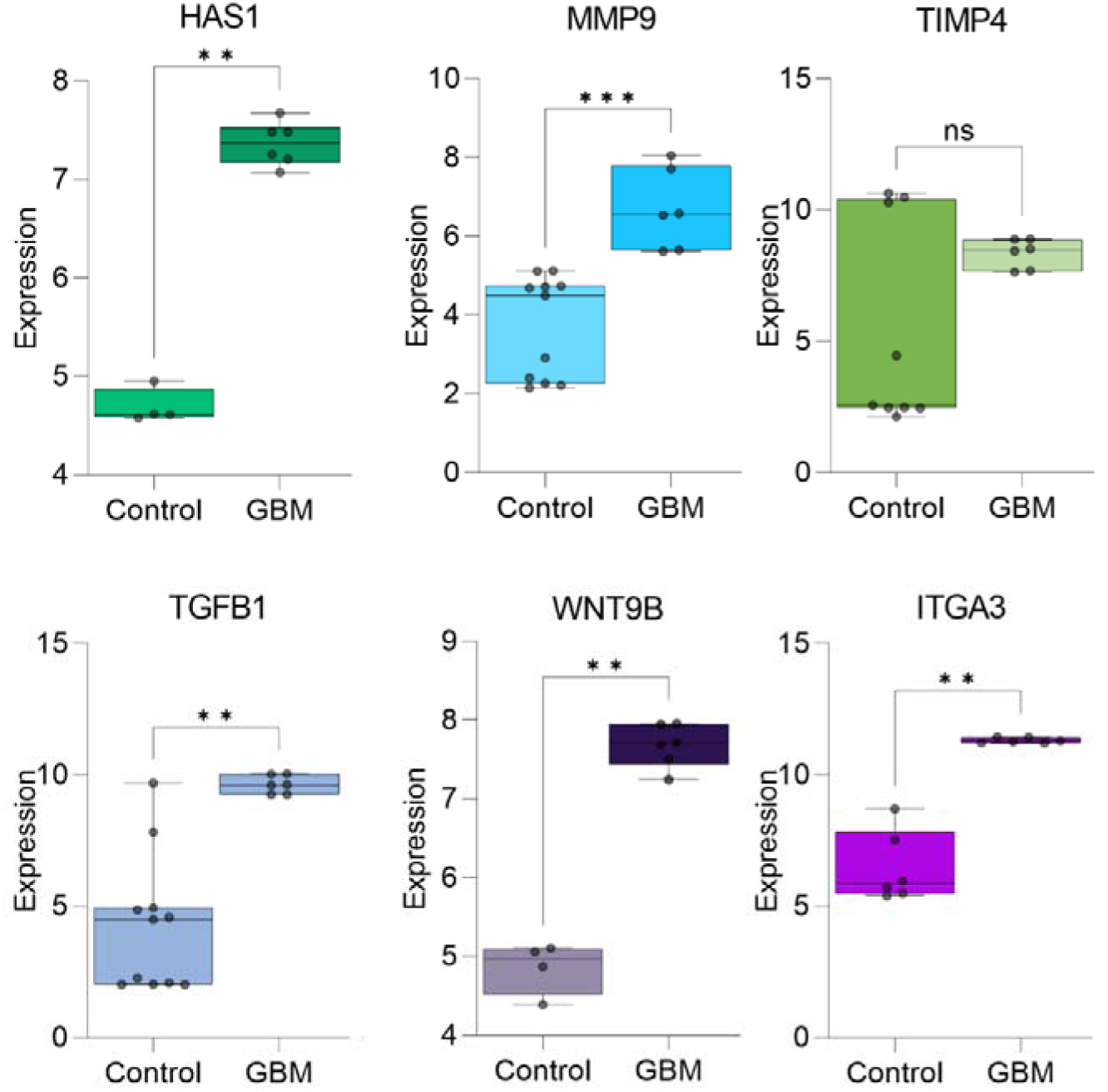
EC exhibit increased expression of ECM-related molecules under the influence of GBM. We examined whether the GBM microenvironment alters the transcript levels of ECM-degrading enzymes, ECM-storing factors, and integrin genes in EC. We utilized data from EndoDB to compare the EC transcriptomes of GBM specimens (N=6) with those of normal brain EC (N=11). Statistical analysis was performed using the D’Agostino-Pearson and Mann[Whitney tests. Our analysis revealed that EC from GBM exhibited increased expression of the enzyme-encoding genes *HAS1* and *MMP9*, as well as the growth factor genes *TGFB1* and *WNT9B*. Additionally, *ITGA3* expression was also increased in GBM EC compared to controls (ns = nonsignificant, ** p < 0.01, *** p < 0.001).

### Enhanced expression of several ECM molecules was associated with a poorer prognosis in patients with GBM

This investigation aimed to assess whether elevated levels of ECM transcripts correlate with the OS of patients with GBM. We analyzed the expression of the same molecules analyzed in the GBM and EC transcriptomes and divided the GBM cohort into high- and low-expression groups. Our findings revealed significant associations (Fig. 6). Specifically, patients with high expression of *COL5A1* had a median survival of 10.41 months, whereas those with low expression had a median survival of 14.92 months (** p = 0.0021). Similarly, for *COL5A3*, the median survival time was 11.24 months for patients with high COL5A3 expression and 15.12 months for patients with low COL5A3 expression (* p = 0.0114). The data for *COL6A1* indicated median survival times of 11.74 and 14.93 months, respectively (* p = 0.0102). For *COL22A1*, the median survival was 12.66 months in the high-expression group and 19.23 months in the low-expression group (*** p = 0.0008). Notably, patients with high and low *COL27A1* expression had median survival times of 11.74 and 16.6 months, respectively (** p = 0.0014). Patients with high FBLN1 expression had a median survival of 11.28 months, whereas those with low FBLN1 expression had a median survival of 16.64 months (** p = 0.0015). For the *FBLN2* high-expression group, survival was 11.84 months, whereas for the FBLN2 low-expression group, survival was 14.04 months (p = 0.1116). In the case of *ITGA3*, the observed survival times were 10.42 months in the high-expression cohort and 14.42 months in the low-expression cohort (** p= 0.0038). For *ITGA5*, the median survival was 10.43 months in the high-expression group and 14.73 months in the low-expression group (* p = 0.0219). In patients with high *ITGB1* expression, the median survival time was 11.01 months, whereas the median survival time was 14.53 months in patients with low ITGB1 expression (* p = 0.0268). Laminin analysis revealed that patients with high *LAMA1* expression had a median survival of 12.76 months, while patients with low LAMA1 expression had a median survival of 17.49 months (* p = 0.0122). Similarly, for *LAMB1*, the medians were 11.24 and 14.93 months, respectively (** p = 0.0083). High expression of *TGFB1* (8.84 versus 14.93, ** p=0.047), *MMP2* (12.62 versus 14.93, * p = 0.0483), *MMP9* (10.82 versus 14.73, * p=0.0149), *TIMP1* (12.62 versus 15.39, * p = 0.0181), *TIMP2* (12.56 versus 17.65, * p = 0.0321) and *HAS1* (11.01 versus 14.73, * p=0.0313) was also correlated with a worse prognosis. Notably, *NCAN* was the sole ECM molecule analyzed that exhibited a better prognosis in the high-expression group, with a median survival of 14.73 months versus 11.84 months (* p = 0.0124). In summary, the increased expression of *COL5A1/3, COL6A1, COL22A1, COL27A1, FBLN1/2, ITGA3/5, ITGB1, LAMA1, LAMB1, HAS1, MMP2/9, TIMP1/2 and TGFB1*, which were also elevated in GBM cells, or mostly in EC with GBM influence, was associated with a negative impact on the OS of patients with GBM.

**Fig. 6.**
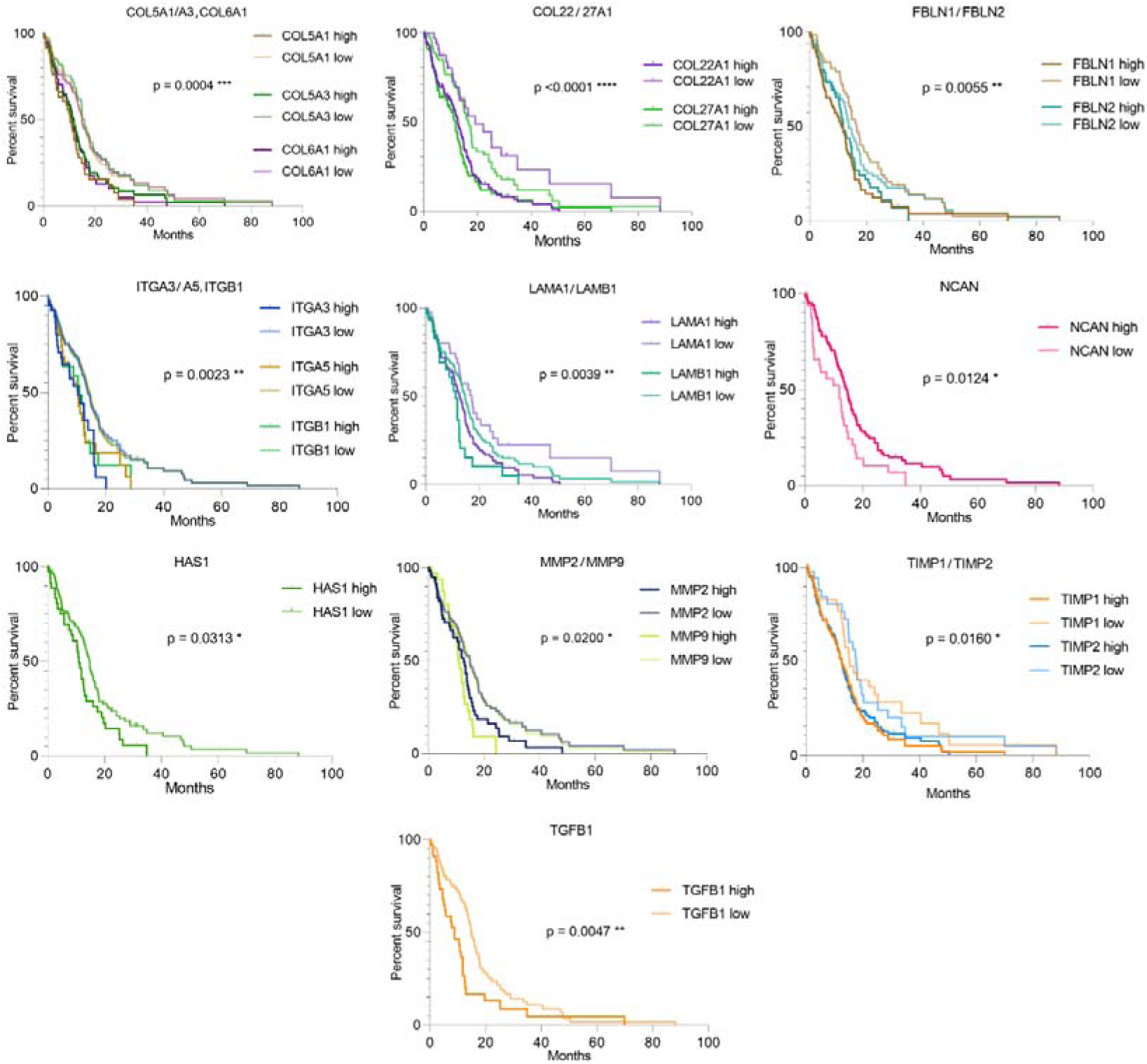
The increased expression of several ECM components was linked to a worse prognosis in GBM patients. We selected the same ECM molecules from the GBM single-cell transcriptome analysis and EC expression analysis to examine their direct influence on the OS of patients with GBM. Using transcriptome data from the Human Protein Atlas and TCGA, we categorized 153 patients into high- and low-expression groups using the lowest log-rank P value obtained. OS curves were constructed using the Mantel[Cox test. Except for *NCAN*, high expression of ECM or ECM-related molecules was associated with poorer prognosis in GBM patients (*p < 0.05, ** p < 0.01, *** p < 0.001, **** p < 0.0001).

## Discussion

GBMs are considered highly angiogenic tumors, and angiogenesis in these tumors is associated with malignancy levels and is negatively correlated with patient survival [48, 49]. During GBM development, tumor cells play an early instructive role in the brain parenchyma. Tumor-instructed stromal cells have differential effects on tumor cell proliferation and migration *in vitro*, indicating reciprocal crosstalk between neoplastic and nonneoplastic cells, such as EC [50, 51]. The composition of the ECM and its stored signaling factors also influence the behavior of GBM cells and GBM stromal cells, affecting GBM angiogenesis, infiltration and aggressiveness [27, 52, 53]. Understanding the complex interplay between the ECM, GBM cells and EC in tumor angiogenesis and tumor aggressiveness is crucial for identifying new biomarkers to improve patient outcomes and for the development of targeted therapies.

We initially standardized a decellularized ECM from GBM [22, 51]. We utilized a gelatin coating to preserve the retention and biophysical properties of the ECM derived from GBM cells [54, 55]. Using this model, our findings indicated that GBM ECM led to a reduction in the number of HBMEC at 24 h and 72 h but not at 48 h. In a similar but slightly different model, Alves et al. (2011) cultured EC from human umbilical vein endothelial cells (HUVEC) on the ECM produced by U373 MG glioma cells. Initially, HUVEC adhered to the immobilized U373 MG matrix; however, a significant proportion of the cells detached and underwent anoikis (50 to 80%) within 24 h compared to cells incubated with control matrices. They showed that surviving EC (20 to 50%) exhibited increased proliferation; however, these EC formed 74-97% fewer tube-like structures *in vitro* than did those grown on nontumoral matrices [27]. It is important to note that both a reduced number of EC and decreased tube formation are critical for creating a hypoxic environment in GBM and subsequently increasing angiogenesis. The dynamics of tumor angiogenesis are very complex and depend on time [22, 56].

Therefore, we investigated the expression of ECM molecules that are increased in GBM compared to normal brain glial cells and could contribute to the decrease in the number of EC. We observed an upregulation of glycoprotein genes, including *COL4A1/2*, *LAMA4*, *LAMB2*, *LAMC1*, *NCAN*, *BVCAN*, and *VCAN,* in GBM cells. Among the ECM-related genes, the *MMP2* and *TIMP1-4* genes showed significantly greater differences between GBM and normal glia. We also observed increased expression of *HAS2*, *TGFB1*, *ITGA3/5* and *ITGB1* in GBM cells. We subsequently examined whether the expression of ECM molecules is altered in EC under the influence of the GBM microenvironment using the EC transcriptome database EndoDB. We observed that the expression levels of most ECM genes (*COL5A3*, *COL6A1*, *COL22A1*, *COL27A1*, *LAMA1*, *LAMB1* and *FBLN1/2*) and ECM-related genes (*HAS1*, *MMP9*, *TGFB1*, *WNT9B* and *ITGA3*) were increased in EC from GBM compared to those from normal brain EC. Therefore, the expression of *TGFB1* and *ITGA3* was greater in both GBM cells and GBM EC than in controls. Additionally, we found a general correlation between increased expression in GBM EC, high expression in GBM specimens, and OS in these patients. For *NCAN*, *MMP2* and *TIMP1/2*, which were more highly expressed in GBM cells than in normal glia, there was also a correlation between their expression in GBM tissue and OS.

Another analysis of the EC transcriptome also demonstrated deregulation of genes involved in angiogenesis and ECM deposition in the vasculature of GBM [57]. We observed enhanced expression of specific collagen subtypes (collagen IV) in GBM cells and collagen V, VI, XXII, and XXVII in EC from GBM tissue. In the nonresponder angiogenesis gene set, *COL4A2* was associated with poor clinical outcomes in bevacizumab-treated patients. This study demonstrated that angiogenesis-associated gene sets are composed of distinct subsets with diverse biological roles and that they represent different clinical responses to antiangiogenic therapy [58]. *COL4A1* and *COL4A2* were also included in a list of 15 genes that composed an immune-related gene signature in gliomas. Based on this gene signature, the OS of patients in the high-risk group was worse than that of patients in the low-risk group [59]. Additionally, immunohistochemistry assays demonstrated that COL6A1 was significantly upregulated in the tumor tissues of high-risk GBM patients. Single-cell RNA sequencing also validated that these malignant cells expressed high levels of *COL6A1* [60]. Increased expression of *COL6A1* and the vascular endothelial molecular marker cluster of differentiation (*CD31)* induced glioma angiogenesis [61]. In another study, the authors developed a novel nomogram based on angiogenesis-related genes, including *COL22A1*, which allowed survival prediction in patients with grade 4 diffuse gliomas [62]. In turn, *COL5A1* has previously been associated with survival patterns in GBM patients (p < 0.0001). Their results indicated that *COL5A1* had the highest ranking, with significantly higher expression levels in GBM tissues than in normal brain tissues. Moreover, *COL5A1* was overexpressed in samples exhibiting aggressive characteristics, including the non-G-CIMP (p < 0.0001), *IDH1* wild-type (p < 0.0001), and unmethylated O^6^-methylguanine-DNA methyltransferase (MGMT) groups (p = 0.008). Regarding the molecular subtype classification of GBM, the expression level of *COL5A1* in the mesenchymal (Mes) subtype was higher than that in the proneural and classical subtypes (p < 0.001) [63]. The expression of *COL5A3* was also correlated with poor survival in GBM patients [64]. Therefore, the expression of some collagen IV, V, VI and XXII subtypes has been associated with increased aggressive characteristics of GBM, such as increased angiogenesis, the GBM mesenchymal subtype and poor survival. Our study is the first to directly correlate high expression of *COL5A3* with a modulated profile of EC in GBM and the first to associate *COL27A1* with poor survival in a GBM cohort.

Laminins are the major components of blood vessel BM [65]. The upregulation of *LAMB1* expression strongly correlated with the downregulation of miR-124-5p. Glioma growth via increased angiogenesis was reversed upon *LAMB1* knockdown [66]. *LAMB1* has been described as a promising candidate biomarker in GBM [67]. Low levels of *LAMB1* have also been associated with a less invasive tumor phenotype and a better prognosis [68]. Endothelial angiogenic phenotypes have been mapped to tumor angiogenesis and included for example, a gene signature associated with vascular BM remodeling, such as *LAMB1*, *LAMA4, COL4A1* and *COL4A2* [69, 70]. In turn, the LAMA1 chain is a component of various laminin isoforms (laminin-1, -2, -6, -8, -10, and -12) [71]. These laminin chains have been previously found to be secreted by glioma cells and play roles in peritumoral infiltration through the adhesion of cancer cells to ECM components and cellular migration [72]. Lower expression of *LAMA1* has been associated with improved survival in GBM patient datasets, whereas *LAMA1* overexpression has been correlated with increased tumor growth in primary GBM [73, 74]. Overall, high *LAMB1* and *LAMA1* expression is associated with glioma growth, infiltration, and poor prognosis. To date, there have been no studies indicating the relationships between *LAMA1, LAMB2* and *LAMC1* expression and angiogenesis in GBM or GBM EC.

Fibulins are elastic fiber–associated proteins. Bioinformatics analysis has demonstrated that angiogenesis-related pathways, which involve differentially expressed fibulin, are increased in low-to high-grade astrocytomas [75]. The overexpression of *FBLN1*, also known as *EFEMP1*, has been shown to eliminate tumor development and suppress angiogenesis, cell proliferation, and *VEGFA* expression. In GBM cells treated with exogenous FBLN1 protein or overexpressed endogenous FBLN1, the level of EGFR was reduced [76]. Additionally, a study involving 95 GBM patients analyzed by qPCR demonstrated that FBLN1 was a favorable prognostic marker for patients with GBM according to Cox regression analysis [76]. On the other hand, *FBLN2* downregulation was also associated with advanced clinical stage in astrocytoma tissues [75]. These apparently conflicting results between our results and this literature may be explained by the size of the analyzed cohorts, our classification of patients into high and low *FBLN1* expression groups, and the inclusion of only wild-type GBM patients in our cohort. However, further studies are necessary to elucidate the relationships between *FBLN1*/2 expression and GBM angiogenesis and prognosis.

Our study identified *TGFB1* as one of the overexpressed growth factors in GBM and GBM EC. Integrative bioinformatic analysis confirmed the key role of TGFB1 as a major interaction module shaping GBM progression [50]. TGFB1 was found to enhance GBM-induced angiogenesis, a process that was impaired by a c-Jun N-terminal kinase (JNK) inhibitor (SP600125) but not by extracellular signal-regulated kinase (ERK), phosphatidylinositol 3-kinase (PI3K), or mitogen-activated protein kinase (MAPK) inhibitors. These findings underscore the critical role of the TGFB1 and JNK pathways in mediating GBM angiogenesis [77, 78]. *TGFB1* expression was also positively associated with aggressive prognosis, as it was higher in the Mes subtype of high-grade glioma (HGG) than in the proneural subtype. Consistent with our results, overall survival analysis of GBM patients revealed that those in the top 50% *TGFB1* expression group survived significantly shorter than those in the bottom 50% TGFB1 expression group. *TGFB1* has emerged as a potential prognostic molecule and a potential signature gene for Mes-subtype HGG [79].

Furthermore, certain WNT variants have been implicated in angiogenesis in GBM [80, 81]. The WNT signaling pathway is necessary for angiogenesis in the CNS and BBB differentiation. β-catenin, a component of the WNT signaling pathway, serves as a marker for proliferating EC in adult GBM [82]. Phosphorylation of β-catenin at Ser675 and activation of the WNT signaling pathway induced the expression of multidrug resistance-associated protein-1 (MRP-1), leading to EC stemness-like activation and chemoresistance [83]. To our knowledge, this study is the first to describe the potential role of WNT9B expression in GBM angiogenesis.

Our findings showed that GBM cells had increased expression of *ITGB1*, *ITGA5* and *ITGA3* and that this increased expression was correlated with poorer survival in GBM patients. *ITGA3* was also overexpressed in GBM EC. Interestingly, an anti-integrin-based approach to inhibit GBM angiogenesis represents an alternative strategy to the traditional focus on anti-VEGF therapies [84, 85]. The overexpression of *ITGA3*/*ITGB1* plays a central role in promoting tube formation in tumor-associated EC in GBM. Blocking α3β1 reduces the sprouting and tube formation of these EC, as well as vessel density, in organotypic cultures of GBM. Consistent with our findings, the *ITGA3* mRNA expression in gliomas of WHO grades II, III and IV increased successively, with significant differences (p < 0.05). Additionally, high expression of *ITGA3* in GBM significantly and independently predicts poor survival in patients with astrocytoma [86]. Similar to *COL6A1*, the *ITGA5* gene was included in a risk prediction model constructed based on differentially expressed angiogenesis-related genes [60], and this risk score could also serve as a promising prognostic predictor [87].

In our study, we found that *HAS1* expression is increased in GBM EC and that *HAS2* expression is increased in GBM cells. HA strongly impacts tumor development and progression by promoting cell proliferation, angiogenesis, chemotherapy resistance, and invasion of surrounding tissues [26, 88]. Tissue microarray data have shown high expression of *HA* and *HAS1*-*3* in malignant tissue of grade II-IV diffusely infiltrating astrocytomas [89]. In addition, HA increases the levels of ceruloplasmin, a protein related to hypoxia, inflammation, and angiogenesis in gliomas [90]. To date, there have been no studies directly relating *HAS1* expression and EC in GBM; however, an increase in *HAS2* expression has been correlated with poor prognosis in GBM patients [90, 91]. In the database utilized in our study, we have no access to *HAS2* expression versus survival data to perform this survival curve analysis.

The highly invasive growth of GBM is influenced by an abnormal profile of subtypes of MMP and its inhibitor TIMP involved in remodeling of the BM and normal vasculature [92–94]. We found that *MMP9* was more highly expressed in GBM EC and that it was correlated with poorer survival in GBM patients. The role of MMP9 in GBM angiogenesis has long been studied [95, 96]. Increased expression of *MMP9* has been correlated with increased angiogenesis [97], and its expression is significantly induced by hypoxia in GBM [98]. The expression of *MMP9* has also been significantly associated with poor prognosis in GBM patients [99]. Interestingly, the predictive value of MMP9 in a randomized phase III trial in patients with newly diagnosed GBM was correlated with the efficacy of bevacizumab, an antiangiogenic drug. Patients with low MMP9 levels had a significant 5.2-month OS benefit with bevacizumab (p = 0.0009). Multivariate analysis revealed a significant interaction between treatment and MMP9 (p = 0.03) for OS [96]. We also found that *MMP2* and *TIMP1-4* were more highly expressed in GBM than in normal glia and that higher expression of *MMP2* and *TIMP1*/*2* was associated with lower survival in patients with these tumors. To our knowledge, this is the first study that directly revealed an association between high expression of *TIMP2* and poor prognosis in GBM patients. Conroy and collaborators (2017) [100] showed that *MMP2*, *MMP9*, *TIMP1* and *TIMP2* were more highly expressed in GBM of the classical-like subtype. Recently, *TIMP1* was identified as one of eight prognosis-related differentially expressed genes in GBM [101]. Consistent with our results, decreased expression of *TIMP1* has been associated with prolonged survival [102].

Finally, elevated expression levels of the lecticans *VCAN*, *BCAN*, and *NCAN* in GBM have been documented [26], along with their distinct regulation of O-glycopeptides compared to normal tissue [103]. Here, *VCAN*, *BCAN* and *NCAN* were more highly expressed in GBM cells than in normal glia, a phenomenon that may have contributed to the reduction in the number of EC. The VCAN V2 isoform has been shown to promote angiogenesis by modulating EC activity and fibronectin expression in U87 GBM cells [28]. Increased levels of *VCAN* [104, 105] and *BCAN* [106] expression have been associated with invasion and prognosis in GBM patients. To date, direct associations between *NCAN* expression, GBM angiogenesis, and patient survival have not been investigated. *NCAN* emerged as the only ECM gene for which patients with high expression levels exhibited improved prognostic outcomes. Therefore, we propose that evaluating the expression of *NCAN* in GBM may be a useful tool for predicting GBM prognosis.

In future studies, the array of ECM and ECM-associated molecules overexpressed in GBM cells or GBM EC identified here may offer promising avenues for novel antiangiogenic strategies against these tumors. Moreover, considering our finding of a correlation between increased ECM expression and poorer prognosis in GBM patients, we propose that assessing this expression profile alongside established WHO2021 markers in patient tumor tissues may be valuable for obtaining prognostic insights. Additionally, targeting the expression of these genes in GBM may increase the survival rate of GBM patients.

## Conclusion

In summary, GBM-derived ECM decreased the total number of HBMEC at 24 h and 72 h. Increased expression of subtypes of collagens, laminins, fibulins, and the ECM-related genes *ITGA3/5*, *ITGB1*, *TGFB1*, *MMP2/9*, *TIMP1/2* and *HAS1* was observed in GBM cells or EC from GBM, and these molecules were directly linked to the survival of GBM patients. This study revealed, for the first time, that the expression of *COL5A3*, *LAMA1*, *LAMB2*, *LAMC1*, *WNT9B*, and *HAS1* was increased in GBM EC or GBM cells compared to controls, identifying these genes as potential targets for antiangiogenic therapies or for further investigation of GBM angiogenesis. Additionally, except for *NCAN*, increased expression of ECM molecules was associated with shorter survival time in patients with GBM. This study also demonstrated that *NCAN* and *COL27A1* are potential new ECM biomarkers for assisting in GBM prognosis. Increased expression of *NCAN* predicts better prognosis, whereas increased expression of *COL27A1* is linked to worse prognosis. Modulating the expression of these genes in GBM could contribute to enhance prognostic outcomes for patients with the condition.

## Data Availability

All data produced in the present study are available upon reasonable request to the authors

## Author contributions

L.C.B. and M.H. performed the *in vitro* assays. G.C.M. performed the bioinformatic analysis. V.F. designed and supervised the study and wrote and edited the manuscript.

## Acknowledgments

The authors express their gratitude to Prof. Vivaldo Moura Neto for providing the laboratory infrastructure and for being a personal source of inspiration.

## References

1. Louis DN, Perry A, Wesseling P, et al (2021) The 2021 WHO classification of tumors of the central nervous system: A summary. Neuro Oncol 23:. 10.1093/neuonc/noab106

2. Brat DJ, Aldape K, Colman H, et al (2018) cIMPACT-NOW update 3: recommended diagnostic criteria for “Diffuse astrocytic glioma, IDH-wildtype, with molecular features of glioblastoma, WHO grade IV.” Acta Neuropathol 136

3. Wen PY, Weller M, Lee EQ, et al (2020) Glioblastoma in adults: A Society for Neuro-Oncology (SNO) and European Society of Neuro-Oncology (EANO) consensus review on current management and future directions. Neuro Oncol 22

4. Takano S, Yamashita T, Ohneda O (2010) Molecular therapeutic targets for glioma angiogenesis. J Oncol

5. De Bock K, Cauwenberghs S, Carmeliet P (2011) Vessel abnormalization: Another hallmark of cancer?. Molecular mechanisms and therapeutic implications. Curr Opin Genet Dev 21

6. Mazzone M, Dettori D, Leite de Oliveira R, et al (2009) Heterozygous Deficiency of PHD2 Restores Tumor Oxygenation and Inhibits Metastasis via Endothelial Normalization. Cell 136:. 10.1016/j.cell.2009.01.020

7. Nagy JA, Chang SH, Shih SC, et al (2010) Heterogeneity of the tumor vasculature. Semin Thromb Hemost 36

8. Folkins C, Shaked Y, Man S, et al (2009) Glioma tumor stem-like cells promote tumor angiogenesis and vasculogenesis via vascular endothelial growth factor and stromal-derived factor 1. Cancer Res 69:. 10.1158/0008-5472. CAN-09-0167

9. Papetti M, Herman IM (2002) Mechanisms of normal and tumor-derived angiogenesis. Am J Physiol Cell Physiol 282

10. Pen A, Moreno MJ, Martin J, Stanimirovic DB (2007) Molecular markers of extracellular matrix remodeling in glioblastoma vessels: Microarray study of laser-captured glioblastoma vessels. Glia 55:. 10.1002/glia.20481

11. Baish JW, Stylianopoulos T, Lanning RM, et al (2011) Scaling rules for diffusive drug delivery in tumor and normal tissues. Proc Natl Acad Sci U S A 108:. 10.1073/pnas.1018154108

12. Stylianopoulos T, Jain RK (2013) Combining two strategies to improve perfusion and drug delivery in solid tumors. Proc Natl Acad Sci U S A 110:. 10.1073/pnas.1318415110

13. Onishi M, Kurozumi K, Ichikawa T, Date I (2013) Mechanisms of tumor development and anti-angiogenic therapy in glioblastoma multiforme. Neurol Med Chir (Tokyo) 53:. 10.2176/nmc.ra2013-0200

14. Stacker SA, Caesar C, Baldwin ME, et al (2001) VEGF-D promotes the metastatic spread of tumor cells via the lymphatics. Nat Med 7:. 10.1038/84635

15. Weathers SP, de Groot J (2015) VEGF manipulation in glioblastoma. ONCOLOGY (United States) 29

16. Pepper MS, Vassalli JD, Orci L, Montesano R (1993) Biphasic effect of transforming growth factor-β1 on in vitro angiogenesis. Exp Cell Res 204:. 10.1006/excr.1993.1043

17. Burghardt I, Schroeder JJ, Weiss T, et al (2021) A tumor-promoting role for soluble TβRIII in glioblastoma. Mol Cell Biochem 476:. 10.1007/s11010-021-04128-y

18. Rempel SA, Dudas S, Ge S, Gutiérrez JA (2000) Identification and localization of the cytokine SDF1 and its receptor, CXC chemokine receptor 4, to regions of necrosis and angiogenesis in human glioblastoma. Clinical Cancer Research 6:

19. Gonçalves TL, de Araújo LP, Pereira Ferrer V (2023) Tamoxifen as a modulator of CXCL12-CXCR4-CXCR7 chemokine axis: A breast cancer and glioblastoma view. Cytokine 170

20. Stratmann A, Risau W, Plate KH (1998) Cell type-specific expression of angiopoietin-1 and angiopoietin-2 suggests a role in glioblastoma angiogenesis. American Journal of Pathology 153:. 10.1016/S0002-9440(10)65733-1

21. Labussière M, Cheneau C, Prahst C, et al (2016) Angiopoietin-2 May Be Involved in the Resistance to Bevacizumab in Recurrent Glioblastoma. Cancer Invest 34:. 10.3109/07357907.2015.1088948

22. Ahir BK, Engelhard HH, Lakka SS (2020) Tumor Development and Angiogenesis in Adult Brain Tumor: Glioblastoma. Mol Neurobiol 57

23. Senger DR, Davis GE (2011) Angiogenesis. Cold Spring Harb Perspect Biol 3:. 10.1101/cshperspect.a005090

24. De Oliveira Rosario LV, Da Rosa BG, Goncalves TL, et al (2020) Glioblastoma factors increase the migration of human brain endothelial cells in vitro by increasing MMP-9/CXCR4 levels. Anticancer Res 40:. 10.21873/anticanres.14244

25. Lakka SS, Rao JS (2008) Antiangiogenic therapy in brain tumors. Expert Rev Neurother 8

26. Ferrer P (2018) Glioma infiltration and extracellular matrix[: key players and modulators. 1–24. 10.1002/glia.23309

27. Alves TR, da Fonseca ACC, Nunes SS, et al (2011) Tenascin-C in the extracellular matrix promotes the selection of highly proliferative and tubulogenesis-defective endothelial cells. Exp Cell Res. 10.1016/j.yexcr.2011.06.006

28. Yang W, Yee AJ (2013) Versican V2 isoform enhances angiogenesis by regulating endothelial cell activities and fibronectin expression. FEBS Lett 587:. 10.1016/j.febslet.2012.11.023

29. Kretschmer M, Rüdiger D, Zahler S (2021) Mechanical aspects of angiogenesis. Cancers (Basel) 13

30. Petrik J, Campbell NE, Kellenberger L, et al (2010) Extracellular matrix proteins and tumor angiogenesis. J Oncol

31. Rupp T, Langlois B, Koczorowska MM, et al (2016) Tenascin-C Orchestrates Glioblastoma Angiogenesis by Modulation of Pro- and Anti-angiogenic Signaling. Cell Rep. 10.1016/j.celrep.2016.11.012

32. Zagzag D, Friedlander DR, Miller DC, et al (1995) Tenascin Expression in Astrocytomas Correlates with Angiogenesis. Cancer Res 55:

33. Behrem S, Žarković K, Eškinja N, Jonjić N (2005) Distribution pattern of tenascin-C in glioblastoma: Correlation with angiogenesis and tumor cell proliferation. Pathology and Oncology Research 11:. 10.1007/BF02893856

34. Mammoto T, Jiang A, Jiang E, et al (2013) Role of collagen matrix in tumor angiogenesis and glioblastoma multiforme progression. American Journal of Pathology 183:. 10.1016/j.ajpath.2013.06.026

35. Yunker CK, Golembieski W, Lemke N, et al (2008) SPARC-induced increase in glioma matrix and decrease in vascularity are associated with reduced VEGF expression and secretion. Int J Cancer 122:. 10.1002/ijc.23450

36. Liu D, Pearlman E, Diaconu E, et al (1996) Expression of hyaluronidase by tumor cells induces angiogenesis in vivo. Proc Natl Acad Sci U S A 93:. 10.1073/pnas.93.15.7832

37. Ljubimova JY, Fujita M, Khazenzon NM, et al (2006) Changes in laminin isoforms associated with brain tumor invasion and angiogenesis. Frontiers in Bioscience 11

38. Faria J, Romão L, Martins S, et al (2006) Interactive properties of human glioblastoma cells with brain neurons in culture and neuronal modulation of glial laminin organization. Differentiation. 10.1111/j.1432-0436.2006.00090.x

39. Stins MF, Badger J, Sik Kim K (2001) Bacterial invasion and transcytosis in transfected human brain microvascular endothelial cells. Microb Pathog 30:. 10.1006/mpat.2000.0406

40. Bhattacharyya D, Hammond AT, Glick BS (2010) High-quality immunofluorescence of cultured cells. Methods Mol Biol 619:. 10.1007/978-1-60327-412-8_24

41. Chaligne R, Gaiti F, Silverbush D, et al (2021) Epigenetic encoding, heritability and plasticity of glioma transcriptional cell states. Nat Genet 53:. 10.1038/s41588-021-00927-7

42. Khan S, Taverna F, Rohlenova K, et al (2019) EndoDB: A database of endothelial cell transcriptomics data. Nucleic Acids Res 47:. 10.1093/nar/gky997

43. Banerjee A, Kim BJ, Carmona EM, et al (2011) Bacterial Pili exploit integrin machinery to promote immune activation and efficient blood[brain barrier penetration. Nat Commun 2:. 10.1038/ncomms1474

44. Ricci-Vitiani L, Pallini R, Biffoni M, et al (2010) Tumor vascularization via endothelial differentiation of glioblastoma stem-like cells. Nature 468:. 10.1038/nature09557

45. Karlsson M, Zhang C, Méar L, et al (2021) A single–cell type transcriptomics map of human tissues. Sci Adv 7:. 10.1126/sciadv.abh2169

46. Cerami E, Gao J, Dogrusoz U, et al (2012) The cBio cancer genomics portal: an open platform for exploring multidimensional cancer genomics data. Cancer Discov 2:401–404. 10.1158/2159-8290. CD-12-0095

47. Gao J, Aksoy BA, Dogrusoz U, et al (2013) Integrative analysis of complex cancer genomics and clinical profiles using the cBioPortal. Sci Signal 6:pl1. 10.1126/scisignal.2004088

48. Zhang Y, Zhang N, Dai B, et al (2008) FoxM1B transcriptionally regulates vascular endothelial growth factor expression and promotes the angiogenesis and growth of glioma cells. Cancer Res 68:. 10.1158/0008-5472. CAN-08-1968

49. Alves TR, Lima FRS, Kahn SA, et al (2011) Glioblastoma cells: A heterogeneous and fatal tumor interacting with the parenchyma. In: Life Sciences. pp 532–539

50. Bougnaud S, Golebiewska A, Oudin A, et al (2016) Molecular crosstalk between tumor and brain parenchyma instructs histopathological features in glioblastoma. Oncotarget 7:. 10.18632/oncotarget.7454

51. Testa E, Palazzo C, Mastrantonio R, Viscomi MT (2022) Dynamic Interactions between Tumor Cells and Brain Microvascular Endothelial Cells in Glioblastoma. Cancers (Basel) 14

52. Dinevska M, Widodo SS, Furst L, et al (2023) Cell signaling activation and extracellular matrix remodeling underpin glioma tumor microenvironment heterogeneity and organization. Cellular Oncology 46:. 10.1007/s13402-022-00763-9

53. Marino S, Menna G, Di Bonaventura R, et al (2023) The Extracellular Matrix in Glioblastomas: A Glance at Its Structural Modifications in Shaping the Tumoral Microenvironment—A Systematic Review. Cancers (Basel) 15

54. Pedron S, Harley BAC (2013) Impact of the biophysical features of a 3D gelatin microenvironment on glioblastoma malignancy. J Biomed Mater Res A 101:. 10.1002/jbm.a.34637

55. Harris GM, Raitman I, Schwarzbauer JE (2018) Cell-derived decellularized extracellular matrices. In: Methods in Cell Biology

56. Lee SY, Park JH, Cho KH, et al (2022) Isolinderalactone inhibits glioblastoma cell supernatant-induced angiogenesis. Oncol Lett 24:. 10.3892/ol.2022.13448

57. Schaffenrath J, Wyss T, He L, et al (2021) Blood[brain barrier alterations in human brain tumors revealed by genome-wide transcriptomic profiling. Neuro Oncol 23:. 10.1093/neuonc/noab022

58. Choi SW, Shin H, Sa JK, et al (2018) Identification of transcriptome signature for predicting clinical response to bevacizumab in recurrent glioblastoma. Cancer Med 7:. 10.1002/cam4.1439

59. Gong X, Liu L, Xiong J, et al (2021) Construction of a Prognostic Gene Signature Associated with Immune Infiltration in Glioma: A Comprehensive Analysis Based on the CGGA. J Oncol 2021:. 10.1155/2021/6620159

60. Wan Z, Zuo X, Wang S, et al (2023) Identification of angiogenesis-related genes signature for predicting survival and its regulatory network in glioblastoma. Cancer Med 12:. 10.1002/cam4.6316

61. Han X, Wang Q, Fang S, et al (2022) P4HA1 Regulates CD31 via COL6A1 in the Transition of Glioblastoma Stem-Like Cells to Tumor Endothelioid Cells. Front Oncol 12:. 10.3389/fonc.2022.836511

62. Liu H, Zeng Z, Sun P (2023) Prognosis and immunoinfiltration analysis of angiogene-related genes in grade 4 diffuse gliomas. Aging 15:9842–9857. 10.18632/aging.205054

63. Tsai HF, Chang YC, Li CH, et al (2021) Type V collagen alpha 1 chain promotes the malignancy of glioblastoma through PPRC1-ESM1 axis activation and extracellular matrix remodeling. Cell Death Discov 7:. 10.1038/s41420-021-00661-3

64. Bassot A, Dragic H, Haddad S Al, et al (2023) Identification of a miRNA multitargeting therapeutic strategy in glioblastoma. Cell Death Dis 14:. 10.1038/s41419-023-06117-z

65. Yousif LF, Di Russo J, Sorokin L (2013) Laminin isoforms in endothelial and perivascular basement membranes. Cell Adh Migr 7

66. Chen Q, Lu G, Cai Y, et al (2014) MiR-124-5p inhibits the growth of high-grade gliomas through posttranscriptional regulation of LAMB1. Neuro Oncol 16:. 10.1093/neuonc/not300

67. Zolotovskaia MA, Kovalenko MA, Tkachev VS, et al (2022) Next-Generation Grade and Survival Expression Biomarkers of Human Gliomas Based on Algorithmically Reconstructed Molecular Pathways. Int J Mol Sci 23:. 10.3390/ijms23137330

68. Virga J, Bognár L, Hortobágyi T, et al (2017) Prognostic Role of the Expression of Invasion-Related Molecules in Glioblastoma. J Neurol Surg A Cent Eur Neurosurg 78:. 10.1055/s-0036-1584920

69. Zhang L, He L, Lugano R, et al (2018) IDH mutation status is associated with distinct vascular gene expression signatures in lower-grade gliomas. Neuro Oncol 20:. 10.1093/neuonc/noy088

70. Xie Y, He L, Lugano R, et al (2021) Key molecular alterations in endothelial cells in human glioblastoma uncovered through single-cell RNA sequencing. JCI Insight 6:. 10.1172/jci.insight.150861

71. Patarroyo M, Tryggvason K, Virtanen I (2002) Laminin isoforms in tumor invasion, angiogenesis and metastasis. Semin Cancer Biol 12:. 10.1016/S1044-579X(02)00023-8

72. Kawataki T, Yamane T, Naganuma H, et al (2007) Laminin isoforms and their integrin receptors in glioma cell migration and invasiveness: Evidence for a role of α5-laminin(s) and α3β1 integrin. Exp Cell Res 313:. 10.1016/j.yexcr.2007.07.038

73. Scrideli CA, Carlotti CG, Okamoto OK, et al (2008) Gene expression profile analysis of primary glioblastomas and nonneoplastic brain tissue: Identification of potential target genes by oligonucleotide microarray and real-time quantitative PCR. J Neurooncol 88:. 10.1007/s11060-008-9579-4

74. Fiscon G, Conte F, Licursi V, et al (2018) Computational identification of specific genes for glioblastoma stem-like cells identity. Sci Rep 8:. 10.1038/s41598-018-26081-5

75. Ren T, Lin S, Wang Z, Shang A (2016) Differential proteomics analysis of low-and high-grade of astrocytoma using iTRAQ quantification. Onco Targets Ther 9:. 10.2147/OTT. S111103

76. Hu Y, Pioli PD, Siegel E, et al (2011) EFEMP1 suppresses malignant glioma growth and exerts its action within the tumor extracellular compartment. Mol Cancer 10:. 10.1186/1476-4598-10-123

77. Yang XJ, Chen GL, Yu SC, et al (2013) TGF-β1 enhances tumor-induced angiogenesis via JNK pathway and macrophage infiltration in an improved zebrafish embryo/xenograft glioma model. Int Immunopharmacol 15:. 10.1016/j.intimp.2012.12.002

78. Han J, Alvarez-Breckenridge CA, Wang QE, Yu J (2015) TGF-β signaling and its targeting for glioma treatment. Am J Cancer Res 5

79. Pan YB, Zhang CH, Wang SQ, et al (2018) Transforming growth factor beta induced (TGFBI) is a potential signature gene for mesenchymal subtype high-grade glioma. J Neurooncol 137:. 10.1007/s11060-017-2729-9

80. McCord M, Mukouyama YS, Gilbert MR, Jackson S (2017) Targeting WNT signaling for multifaceted glioblastoma therapy. Front Cell Neurosci 11:. 10.3389/fncel.2017.00318

81. Riganti C, Salaroglio IC, Pinzòn-Daza ML, et al (2014) Temozolomide downregulates P-glycoprotein in human blood[brain barrier cells by disrupting Wnt3 signaling. Cellular and Molecular Life Sciences 71:. 10.1007/s00018-013-1397-y

82. Manoranjan B, Provias JP (2022) β-Catenin marks proliferating endothelial cells in glioblastoma. Journal of Clinical Neuroscience 98:. 10.1016/j.jocn.2022.02.018

83. Huang M, Zhang D, Wu JY, et al (2020) Wnt-mediated endothelial transformation into mesenchymal stem cell–like cells induces chemoresistance in glioblastoma. Sci Transl Med 12:. 10.1126/scitranslmed.aay7522

84. Seystahl K, Weller M (2012) Is there a world beyond bevacizumab in targeting angiogenesis in glioblastoma? Expert Opin Investig Drugs 21

85. Paolillo M, Serra M, Schinelli S (2016) Integrins in glioblastoma: Still an attractive target? Pharmacol Res 113

86. Xiang Z, Zhongwei W, Haigang C, et al (2022) Integrin alpha 3 expression in glioma and its prognostic value in glioma patients. Chinese Journal of Neuromedicine 21:. 10.3760/cma.j.cn115354-20211012-00654

87. Wang G, Hu JQ, Liu JY, Zhang XM (2022) Angiogenesis-Related Gene Signature-Derived Risk Score for Glioblastoma: Prospects for Predicting Prognosis and Immune Heterogeneity in Glioblastoma. Front Cell Dev Biol 10:. 10.3389/fcell.2022.778286

88. Jong A, Wu CH, Shackleford GM, et al (2008) Involvement of human CD44 during *Cryptococcus neoformans* infection of brain microvascular endothelial cells. Cell Microbiol 10:. 10.1111/j.1462-5822.2008.01128.x

89. Valkonen M, Haapasalo H, Rilla K, et al (2018) Elevated expression of hyaluronan synthase 2 associates with decreased survival in diffusely infiltrating astrocytomas. BMC Cancer 18:. 10.1186/s12885-018-4569-1

90. Pibuel MA, Poodts D, Díaz M, et al (2021) The scrambled story between hyaluronan and glioblastoma. Journal of Biological Chemistry 296

91. Bao X, Ran J, Kong C, et al (2023) Pancancer analysis reveals the potential of hyaluronate synthase as therapeutic targets in human tumors. Heliyon 9:. 10.1016/j.heliyon 2023.e19112

92. Saxena A, Robertson JT, Kufta C, et al (1995) Increased expression of gelatinase A and TIMP-2 in primary human glioblastomas. Int J Oncol 7:. 10.3892/ijo.7.3.469

93. Zerrouqi A, Pyrzynska B, Febbraio M, et al (2012) P14ARF inhibits human glioblastoma-induced angiogenesis by upregulating the expression of TIMP3. Journal of Clinical Investigation 122:. 10.1172/JCI38596

94. Pullen NA, Anand M, Cooper PS, Fillmore HL (2012) Matrix metalloproteinase-1 expression enhances tumorigenicity as well as tumor-related angiogenesis and is inversely associated with TIMP-4 expression in a model of glioblastoma. J Neurooncol 106:. 10.1007/s11060-011-0691-5

95. Thorns V, Walter GF, Thorns C (2003) Expression of MMP-2, MMP-7, MMP-9, MMP-10 and MMP-11 in Human Astrocytic and Oligodendroglial Gliomas. Anticancer Res 23:

96. Jiguet-Jiglaire C, Boissonneau S, Denicolai E, et al (2022) Plasmatic MMP9 released from tumor-infiltrating neutrophils is predictive for bevacizumab efficacy in glioblastoma patients: an AVAglio ancillary study. Acta Neuropathol Commun 10:. 10.1186/s40478-021-01305-4

97. Annabi B, Lachambre MP, Plouffe K, et al (2009) Propranolol adrenergic blockade inhibits human brain endothelial cells tubulogenesis and matrix metalloproteinase-9 secretion. Pharmacol Res. 10.1016/j.phrs.2009.05.005

98. Emara M, Allalunis-Turner J (2014) Effect of hypoxia on angiogenesis related factors in glioblastoma cells. Oncol Rep 31:. 10.3892/or.2014.3037

99. Sun J, Zhao B, Du K, Liu P (2019) TRAF6 correlated to invasion and poor prognosis of glioblastoma by elevating MMP9 expression. Neuroreport 30:. 10.1097/WNR.0000000000001171

100. Conroy S, Wagemakers M, Walenkamp AME, et al (2017) Novel insights into vascularization patterns and angiogenic factors in glioblastoma subclasses. J Neurooncol 131:. 10.1007/s11060-016-2269-8

101. Dang HH, Ta HDK, Nguyen TTT, et al (2023) Identification of a Novel Eight-Gene Risk Model for Predicting Survival in Glioblastoma: A Comprehensive Bioinformatic Analysis. Cancers (Basel) 15:. 10.3390/cancers15153899

102. Jovčevska I (2019) Genetic secrets of long-term glioblastoma survivors. Bosn J Basic Med Sci 19

103. Sethi MK, Downs M, Shao C, et al (2022) In-Depth Matrisome and Glycoproteomic Analysis of Human Brain Glioblastoma Versus Control Tissue. Molecular and Cellular Proteomics 21:. 10.1016/j.mcpro.2022.100216

104. Onken J, Moeckel S, Leukel P, et al (2014) Versican isoform V1 regulates proliferation and migration in high-grade gliomas. J Neurooncol 120:. 10.1007/s11060-014-1545-8

105. László S, József V, Tibor H, et al (2020) Prognostic significance of invasion in glioblastoma. Ideggyogy Sz 73:. 10.18071/isz.73.0317

106. Virga J, Szivos L, Hortobágyi T, et al (2019) Extracellular matrix differences in glioblastoma patients with different prognoses. Oncol Lett 17:. 10.3892/ol.2018.9649

